# Prediction of preeclampsia by a combination of cell-free fetal hemoglobin, cell-free fetal DNA and maternal obstetric history

**DOI:** 10.1101/2025.03.30.25324756

**Authors:** Leif Kofoed Nielsen, Tanja Roien Jakobsen, Sharri Junadi Mortensen, Steen Ladelund, Klaus Rieneck, Frederik Banch Clausen, Jens René Bundgaard, Morten Hanefeld Dziegiel

**Author notes:** **Corresponding author**: Leif Kofoed Nielsen Biomedical Laboratory Science Department of Technology Faculty of Health, University College Copenhagen Sigurdsgade 26, 2200 Copenhagen Denmark.

## Abstract

**Background:** The aim of the study was to investigate the combination of cell-free fetal hemoglobin (cfHbF), total cell-free hemoglobin (tcfHb), cell-free fetal DNA (cffDNA), Pregnancy Associated Plasma Protein A (PAPP-A) and maternal obstetric history as potential markers in first and second trimester, for prediction of preeclampsia (PE) in asymptomatic pregnant women.

**Methods:** This was a retrospective case-control study of 589 pregnant women (560 in the control group and 29 with PE in the case group). We used routine blood samples used in screening for chromosomal anomalies and rhesus immunization to analyze the levels of cfHbF, tcfHb, and PAPP-A in gestational weeks 8–14 and levels of cfHbF, tcfHb, and cffDNA in gestational weeks 23–28.

**Results:** In first-trimester samples, there was no statistically significant difference between controls, mild PE, and severe PE regarding levels of PAPP-A, cfHbF, tcfHb or cfHbF/tcfHb ratio. In second-trimester samples, cfHbF and cffDNA levels were significantly higher in PE cases than in controls. The odds ratio (OR) for developing PE was 4.28 (95%CI: 1.85–9.89) given a cfHbF>90^th^ centile (p<0.001), and 6.55 (95%CI: 2.61–16.46) given a cffDNA level>90^th^ centile (p<0.001). When adjusting for BMI and previous history of PE in a logistic regression analysis, only cffDNA and BMI remained statistically significant (p< 0.001).

**Conclusion:** We found a significant association between PE and high levels of cfHbF and cffDNA in second-trimester samples, but no statistically significant difference in first-trimester samples were observed.

## Introduction

Preeclampsia (PE) is among the most serious pregnancy complications with high morbidity and mortality rates. A substantial 4.6% of all pregnancies are complicated by PE^1^. There has been a great interest in investigating the prediction of PE before onset of clinical symptoms ^2^. The purpose of early prediction is to focus surveillance during pregnancy for mothers and fetuses at risk and initiate possible preventive measures ^3^. Increased surveillance holds the promise of reducing mortality and morbidity rates by initiating prophylactic measures, planning proper timing and place of delivery to enhance chances of fetal and maternal survival.

PE is a syndrome diagnosed after gestational age (GA) 20 weeks in prior normotensive women. The clinical definition of PE is based on cardinal symptoms as high blood pressure (>140/90mm Hg) and proteinuria (>0.3 g/24 hours) ^4^. The pathogenesis of PE is associated with incomplete re-modeling of the spiral arteries failing to transform into low resistance vessels ^5^. The physiological consequence is high vascular resistance which diminishes blood flow to the placenta and consequently to the growing fetus ^6^. The underlying cause of PE is assumed to be an exaggerated immunological response to pregnancy and especially lack of tolerance to the allogeneic fetus with ensuing inflammation and endothelial dysfunction causing the maternal symptoms of PE ^7^.

Several biochemical markers have been tested as screening markers to predict PE prior to onset of clinical symptoms ^8–11^. One potential marker is cell-free fetal hemoglobin (cfHbF) in maternal plasma, which hypothetically would be elevated in case of reduced oxygenation to the developing placenta ^12,13^. Cell-free fetal DNA (cffDNA) in maternal plasma is a normal finding during pregnancy. The cffDNA originates from the physiological turnover of the syncytiotrophoblast. We and others have previously reported increased concentrations of cffDNA and Pregnancy Associated Plasma Protein A (PAPP-A) in maternal plasma preceding development of PE ^14–19^.

### Objectives

The aim of the present study was to assess the combination of cfHbF, cffDNA, PAPP-A and maternal characteristics in improving diagnostic and prediction of PE at two timepoints, GA 8-14 and 23-28 weeks.

## Methods

This was a retrospective case-control study in the Capital Region of Denmark during a two-year period. In Denmark, RHD negative pregnant women are offered a routine fetal antenatal *RHD* screening at GA 25 weeks. Our cohort consisted of RhD negative women who were tested at GA 25 along with first-trimester risk assessment at GA 8+0 to 13+6 weeks ^20^.

The criteria for inclusion was as following: 1) participation in the first trimester risk assessment tests, and 2) participation in the antenatal *RHD* screening performed at GA 23–28 weeks (referred to as GA 25 weeks), and 3) singleton pregnancy. Women without PE or spontaneous delivery before GA 37 weeks were included as controls. PE was diagnosed as new onset hypertension and either proteinuria and/or end-organ dysfunction. PE was grouped into either mild or severe, with severe cases being defined by severe hypertension (systolic BP ≥160 mmHg and/or diastolic BP ≥ 110 mmHg), Intrauterine Growth Retardation (IUGR) and onset of PE prior to GA 34 weeks. The rest were considered mild cases. Women who had been diagnosed with gestational hypertension, proteinuria without hypertension and essential hypertension were excluded from the analysis.

### Biomarkers

Pregnant women in Denmark are offered a first-trimester risk assessment at GA 8+0 to 13+6 weeks, in which PAPP-A and free β-hCG are analyzed. Surplus serum from these samples was stored at −80°C. The logistics of these samples have previously been described ^14,15,21^. Surplus plasma from the samples at GA 25 weeks was stored consecutively at −25°C at the date of analysis.

Total cell-free hemoglobin (tcfHb) and cfHbF were quantified in all samples by enzyme-linked immunosorbent assay (ELISA) ^22^. CffDNA was quantified in samples from the antenatal *RHD* screening performed in GA 25 weeks as described by Clausen et al. ^20^; PAPP-A was measured in weeks 8+0 to 13+6.

### Competitive ELISA for quantification of cfHbF

An indirect competitive ELISA was used for the quantification of cfHbF using 96-well microtiter plates (Nunc, Roskilde, Denmark). Sample diluted in PBS+1%BSA was mixed with anti-HbF (WBAC HbF1, Trillium Diagnostics, Brewer, Maine) diluted 1:7,500 in PBS+1%BSA and incubated for one hour at room temperature. This mixture was transferred to wells in a microtiter plate (MaxiSorp, Nunc, Denmark) coated with HbF (10 µg/mL) and incubated for one hour. After five wash steps we added alkaline phosphatase conjugated rabbit anti-mouse IgG (DAKO, Glostrup, Denmark) diluted 1:500 in PBS+1%BSA.

After a final washing, substrate (p-nitrophenyl phosphate, 1 mg/mL in 1 M diethanolamine containing 0.5 mM MgCl2, pH 9.8) was added and after approximately 15 minutes, the absorbance was read at 405 nm with 492 nm as reference. The limit of detection was 16 ng/mL, with an inter assay coefficient of variation (CV) of 21.1% and an intra assay CV of 9.1%.

### Capture ELISA for quantification of tcfHb

MaxiSorp plates were coated with affinity purified sheep anti-human Hb (Bethyl Laboratories, Montgomery, TX, US) diluted 1:200 in PBS. After five times of washing, the wells were blocked with PBS+1%BSA. Samples were diluted in PBS+1%BSA and applied to the wells. After one-hour incubation at room temperature the wells were washed five times. Bound Hb was detected with Horse Radish Peroxidase (HRP)–conjugated sheep anti-human Hb diluted 1:60,000 (Bethyl Laboratories). After one-hour incubation the wells were washed five times and substrate was added (TMB One Component HRP Microwell Substrate, Bethyl Laboratories). The reaction was stopped by adding 0.2 M H_2_SO_4_ and measured at 450 nm with 620 nm as reference. The limit of detection was 4 ng/mL, with an inter assay coefficient of variation (CV) of 25.7% and an intra assay CV of 4.4%. The ratio of cfHbF to tcfHb was calculated and is referred to as cfHbF/tcfHb ratio.

### CffDNA

The concentration of cffDNA was measured as previously described^15,21^. The 90^th^ centile, which we had set at 122 genome equivalents per milliliter (geq/mL) in an independent cohort^15^, was used to define the highest levels.

### PAPP-A

PAPP-A was measured on freshly collected serum samples as part of the routine antenatal care in weeks 8+0 to 13+6 using an immunology analyzer (Kryptor, Brahms AG, Berlin, Germany) and converted into multiples of the median (MoM) after adjusting for maternal age, weight, smoking, ethnicity, mode of conception, and GA using Astraia software (Astraia Software GmbH, Germany).

### Statistical methods

Continuous data were reported as medians with interquartile ranges (IQR) and compared across groups with the Kruskall Wallis test. Discrete variables were reported as counts and percentages on the form n (%) and compared across groups using Pearson’s Chi-squared test or Fisher’s exact test where appropriate due to data sparseness. As data were sampled according to a case-control protocol, all multiple analyses were logistic regressions reported as Odds Ratios (OR) with 95% confidence intervals (CI). We calculated the 90^th^ percentile to investigate whether the highest levels of cfHbF would be a more sensitive marker. The predictive ability of the variables was examined through use of Receiver Operating Curves (ROC) and summarized as the Area Under the Curve (AUC) with 95% CI. Data management was performed primarily in SAS, and all statistical analyses were done using R Core team (2017).

### Ethics

The study was approved by the Danish Data Protection Agency (ref. 2014-41-2737) and by the Danish Research Ethics Committee (ref. H-3-2012-040). Data regarding previous maternal history and outcome on neonates were obtained from the National Patient Registry (LPR), the Danish Birth Registry (MFR), and the Danish Fetal Medicine Database (jr.nr. FSEID-00000488).

## Results

There were 589 pregnant women included in this study, with 560 women in the control group (no preeclampsia) and 29 women with PE in the case group. In the PE group, 18 (62%) women were diagnosed with mild PE and 11 (38%) women with severe PE.

Maternal characteristics are outlined in **Table 1** according to severity of PE. Women developing PE had significantly higher pre-pregnancy BMI than controls (p=0.003) and women developing mild PE had a significantly higher prevalence of PE in prior pregnancies (p=0.025). Women with PE delivered at an earlier GA than the controls (GA 38 weeks vs. 40 weeks, p<0.001) and had neonates with correspondingly lower birth weight (2,729 g vs. 3,485 g, p=0.003). The incidence of children born small for gestational age was also higher among women developing PE (28% vs. 5%, p<0.001). No difference in placental weight was found.

**Table 1.**
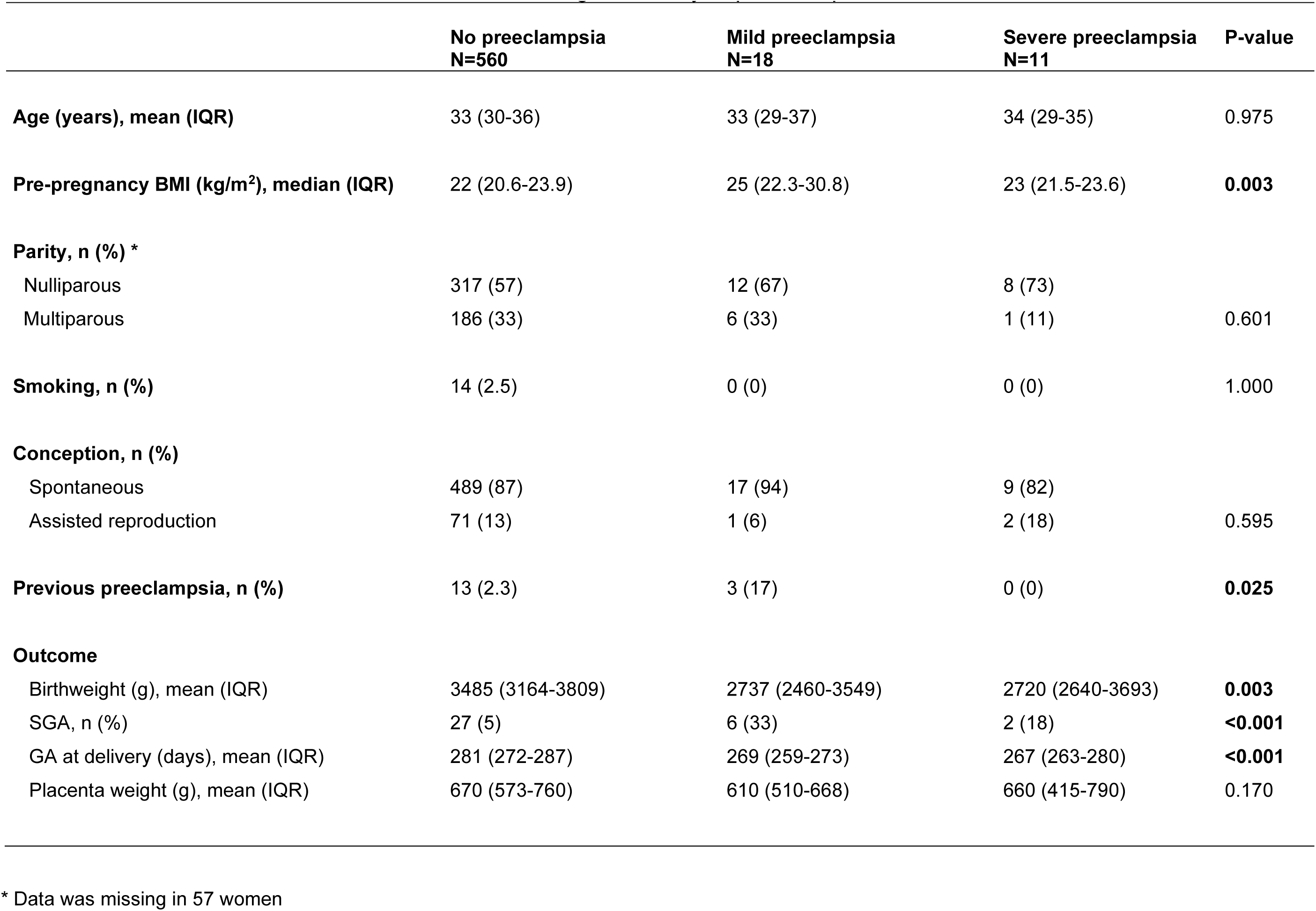
Maternal characteristics and outcome according to severity of preeclampsia

Biochemical markers quantified in first- and second trimester are outlined in **Table 2**. In first-trimester samples, there was no statistically significant difference between controls, mild PE, and severe PE regarding levels of PAPP-A, cfHbF, tcfHb or cfHbF/tcfHb ratio. Overall, there was a higher concentration of both cfHbF and tcfHb levels in second-trimester samples compared with first-trimester samples. The median concentration of cfHbF was significantly higher in cases of PE compared with healthy controls (p=0.007). We found that 45% of women developing severe PE had cfHbF levels above the 90^th^ centile (>28,000 ng/mL, p < 0.001). In contrast, the level of second trimester tcfHb was lower in both mild and severe PE compared with controls (p=0.027). The cfHbF/tcfHb ratio was higher in both mild and severe PE, with the highest ratio observed for severe PE (55.4 vs. 11.4 vs. 8.98, p= 0.008). The median concentration of cffDNA was also significantly higher in severe PE (237 geq/ml) than in both mild PE (103 geq/ml) and in controls (68.5 geq/ml), p=0.002.

**Table 2.**
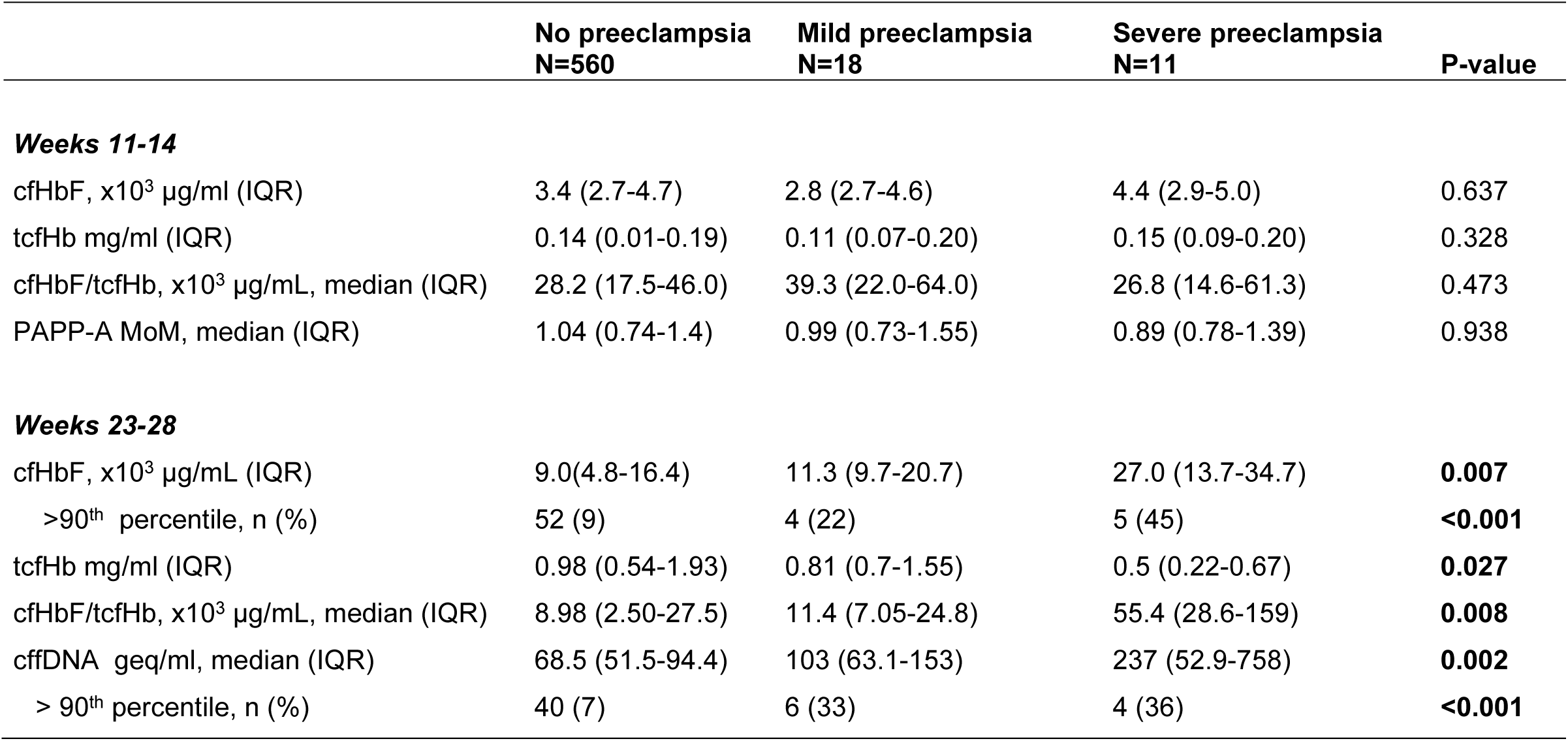
Biochemical markers in weeks 8-14 and 23-28 according to severity of preeclampsia

In an univariate analysis, we found an OR of 4.28 (95% CI: 1.85-9.89) for developing PE given a cfHbF level above the 90^th^ centile (p< 0.001) and an OR of 6.55 (95% CI: 2.61-16.46) for developing PE given a cffDNA level above the 90^th^ centile (p< 0.001). When adjusting for previous history of PE in a combined multivariate regression, we found that only cffDNA and BMI remained significant markers for PE (p< 0.001) (**Table 3**).

**Table 3.** Regression analyses output for the prediction of preeclampsia.

|  | Crude OR (95% CI) | P-value | Multivariate OR (95% CI)* | P-value |
| --- | --- | --- | --- | --- |
| cfHbF > 90th centile | 4.28 (1.9-9.9) | **0.007** | 2.38 (0.7-8.4) | 0.178 |
| cffDNA > 90th centile | 6.58 (2.7-16.2) | **< 0.001** | 11.2 (3.8-33.1) | **< 0.001** |
| BMI | 1.15 (1.1-1.2) | **< 0.001** | 1.21 (1.1-1.3) | **< 0.001** |
| Previous preeclampsia | 4.74 (1.3-17.8) | **< 0.001** | 4.9 (0.8-31.3) | 0.091 |

\* Adjusted for BMI and previous history of PE

ROC curves were drawn for all cases of PE (**Figure 1**) and for cases with severe PE only (**Figure 2**). We found that cfHbF is equal to cffDNA in the prediction of PE, in general, while adding BMI and prior history only increases the sensitivity by 4 and 6 %, respectively (**Figure 1**). For the prediction of severe PE, cfHbF is performing better with an AUC of 0.74 compared to 0.61 for cffDNA. Combining the two markers with BMI and prior history increases the AUC to 0.77, representing an increase in performance of 4% compared to cfHbF alone, but an increase in performance of 26.2% compared to cffDNA as the only marker (**Figure 2**).

**Figure 1.**
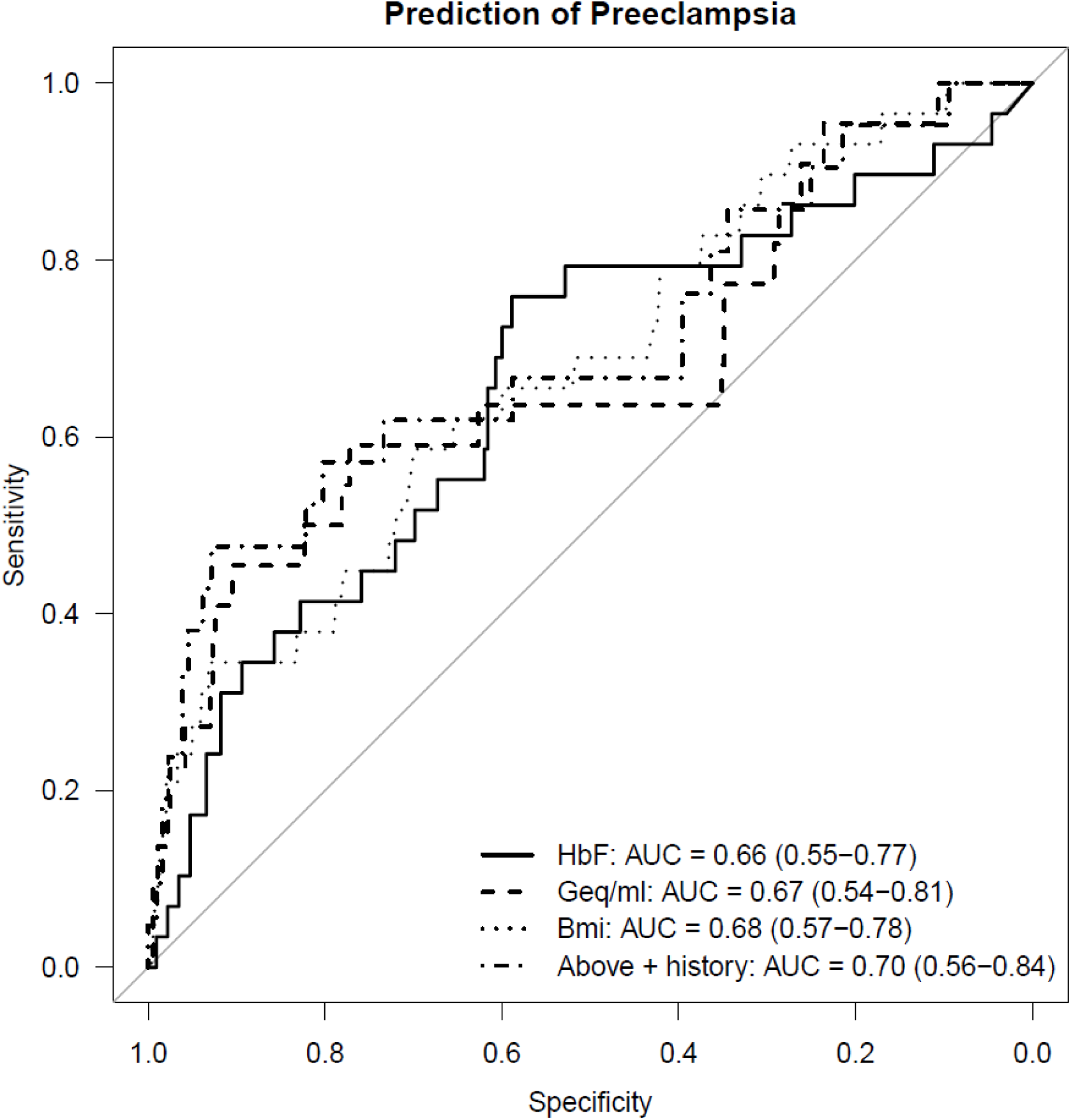
ROC curve for preeclampsia. The curve shows that the AUĆs for cffDNA, HbF and maternal BMI are similar in the prediction of preeclampsia and that the additon of maternal obstetric history to the biomarkers does not increase the detection rate to a clinical useful level.

**Figure 2.**
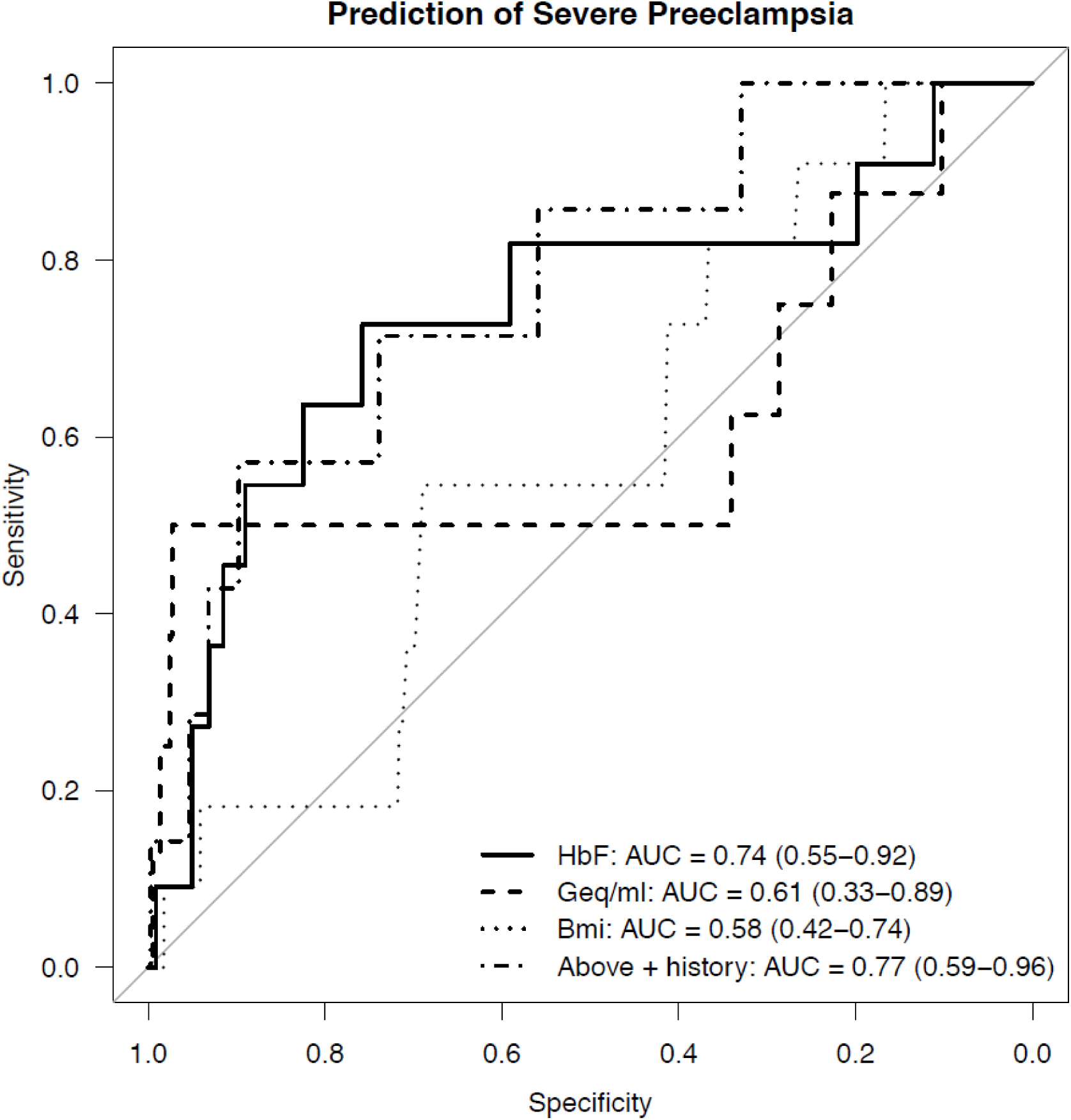
ROC curve for severe preeclampsia. The curve shows that using HbF as a single biomarker is more predictive than a combination of all single markers though the addition of maternal obstetric history only slightly increases AUC.

## Discussion

### Main findings

This study assessed the concentrations of cfHbF as a first and second-trimester marker and cffDNA as a second-trimester marker for PE, with each marker evaluated separately, then combined, and lastly in combination with readily available clinical information from maternal obstetric history. In second-trimester samples, we found a highly significant association between cfHbF, cfHbF/tcfHb ratio, and cffDNA and subsequent PE. When adjusting for BMI and previous history of PE, we found that only cffDNA and BMI remained significant markers for PE. In first-trimester samples, there was no statistically significant difference between controls, mild PE, and severe PE regarding levels of PAPP-A, cfHbF, tcfHb or cfHbF/tcfHb ratio.

### First trimester markers

In the first-trimester samples, we found no significant difference in cfHbF levels between women developing PE and controls. This is in contrast to the study by Anderson et al. who reported a significantly increased cfHbF concentration in the first-trimester in women who later developed PE ^12^. The difference between the proportions of cases to controls in the two studies may explain the dissimilar results. They had a high proportion (62.5%) of PE cases in their case-control study, 60 cases to 36 controls, whereas our study included 29 cases to 560 controls (5%). Furthermore, their patients were recruited as part of an on-going prospective study on serum and ultrasound markers at the first-trimester routine visit, whereas we retrospectively included all women consecutively diagnosed with PE and healthy controls in a three-year period. Moreover, our first-trimester samples were collected at a median GA of 11 weeks, in contrast to their study with a median of 14 weeks for PE cases and 13 weeks for controls ^12^. As fetal utilization of hemoglobin genes shifts from embryonic to fetal in GA weeks 10 to 12, sampling at a too early GA could be counterproductive when measuring fetal hemoglobin, as the gene product is too scarce at this time in pregnancy.

### Second trimester markers

In second trimester samples, we found a significantly elevated concentration of cfHbF, tcfHb as well as cfHbF/tcfHb ratio in women with subsequent PE. The elevation in cfHbF and the cfHbF/tcfHb ratio was especially pronounced for severe PE, which supports the hypothesis that hypoxic events are involved in PE^12^. Similarly, 45% of the women developing PE were found in the group of women with cfHbF above the 90^th^ centile.

High levels of cffDNA were significantly associated with developing PE. In the multivariate analysis, where we adjusted for BMI and prior PE, only cffDNA and BMI remained significant markers for PE. A possible explanation could be that cfHbF is confounded by both BMI and cffDNA.

The inter- and intraassay coefficient of variation of our cfHbF assay was relatively high and may have reduced the detection of women developing PE. Nevertheless, we were able to detect a significant difference in cfHbF concentration and in cfHbF/tcfHb ratio between women developing PE, and controls.

### Preeclampsia and biomarkers

PE is a complex disease predominantly diagnosed in the second or third trimester. One hypothesis is that the underlying cause of PE may be attributed to the lack of tolerance from the maternal immune system towards fetal cells ^23,24^. This impedes proper development of the spiral arteries into low resistance vessels thus hindering a sufficient supply of oxygenated blood to the fetus ^25^. This results in fetal hypoxia and inflammatory endothelial damage with symptoms of multiorgan involvement. cffDNA is released from the placenta as part of the physiological cell turnover in the syncytiotrophoblast. Increased release is seen in preeclamptic women possibly due to the reduced oxygenation and increased apoptosis and necrosis of the syncytiotrophoblast ^26^. It has been proposed that excessive amounts of cffDNA originating from the placenta may enhance the pathogenic process as hypomethylated placental DNA resembles prokaryotic DNA and is assumed to bind preferentially to maternal TLR9 receptors thus inducing a pro-inflammatory response, among other mediators, through activation of IL6 ^27,28^.

We found that a combination of both second-trimester markers, cfHbF and cffDNA, along with maternal obstetric history predicts PE with a sensitivity of approximately 68% at a FPR of 10%. Although the combination of the different markers did increase the AUC, it was not significant. Combining cfHbF, cffDNA, and maternal history did not improve the prediction of PE to a level that is clinically useful. High levels of cffDNA and cfHbF were associated with PE. No single marker has yet proven clinically useful in the prediction of PE. Several studies have tried different combinations of biochemical and biophysical markers and maternal characteristics ^29,30^. Screening in early pregnancy is crucial if proper prophylaxis is to be initiated. Treatment with aspirin before 16 weeks GA, has shown to reduce the rate of PE among women at high risk due to prior history of PE, essential hypertension, and/or connective tissue diseases ^31,32^.

### Limitations

This study is limited by a small number of cases. Furthermore, the timepoints for screening in this study are not appropriate in regard to potentially initiating treatment with aspirin, as first-trimester markers could not identify women at risk for PE and the second-trimester markers could be too late (week 23-28) in regard to initiating treatment, which is why screening at a more optimal time in gestation may be warranted.

### Conclusion

In the present study, we detected women at risk of PE using second-trimester blood samples with the following markers: cfHbF, tcfHb and cffDNA. When adjusted for BMI and previous history of PE, cffDNA and BMI remained significant markers for PE. Larger studies are warranted to verify these markers.

## Data Availability

Data is available on reasonable request.

## Acknowledgments

We are grateful to Charlotte Lajer, Haris Dizdarevic, Franziska Larsen, Janne Amstrup Møller and Betina Poulsen for expert technical assistance. This project was part of a PhD scholarship and funding was received from Rigshospitalet, Copenhagen University Hospital, The Aase and Einar Danielsen Foundation, the Lundbeck Foundation and Arvid Nilssons Foundation.

